# Interaction between *APOE4* status and lifestyle on brain and cognitive outcomes in cognitively unimpaired older adults

**DOI:** 10.1101/2023.07.17.23292755

**Authors:** Francesca Felisatti, Léa Chauveau, Natalie L Marchant, Fabienne Collette, Harriet Demnitz-King, Tim Whitfield, Florence Requier, Florence Mézenge, Brigitte Landeau, Cassandre Palix, Sacha Haudry, Marion Delarue, Vincent de la Sayette, Denis Vivien, Géraldine Poisnel, Robin de Flores, Gaël Chételat, Julie Gonneaud, MEDIT-AGEING Research Group

**Affiliations:** Normandy University, UNICAEN, INSERM, U1237, PhIND “Physiopathology and Imaging of Neurological Disorders”, Institut Blood and Brain @ Caen-Normandie, Cyceron, 14000 Caen, France; Division of Psychiatry, University College London, London, United Kingdom; GIGA CRC In vivo Imaging, University of Liège, Liège, Belgium; Service de Neurologie, CHU de Caen, Caen, France; Département de Recherche Clinique, CHU Caen-Normandie, Caen, France

**Keywords:** Reserve, aging, Alzheimer’s disease, multimodal neuroimaging, cognition

## Abstract

**INTRODUCTION:** *APOE4* genotype and lifestyle have been associated with Alzheimer’s disease (AD) risk, but how they interact on neuroimaging and cognitive markers of aging and AD remains unclear.

**METHODS:** In 135 cognitively unimpaired older adults from the baseline Age-Well trial, we investigated the interaction between *APOE*4 status and cognitive activity, diet and physical activity on cognition and neuroimaging markers of neurodegeneration and amyloid.

**RESULTS:** Higher cognitive activity correlated with lower medial temporal lobe (MTL) volume and perfusion in *APOE4*-carriers, but increased cognitive performance irrespective of *APOE4* status. Higher adherence to the Mediterranean diet correlated with higher MTL metabolism and attention scores in *APOE4-*carriers. Conversely, physical activity correlated with higher MTL perfusion and lower amyloid in *APOE4*-non-carriers only.

**DISCUSSION:** Genetics and lifestyle factors act through different mechanisms to help *APOE4*-carriers resist or cope with brain alterations and postpone cognitive decline. Our results support the need of personalized lifestyle-based interventions for AD.

**Trial Registration Information:** EudraCT: 2016-002441-36; IDRCB: 2016-A01767-44; ClinicalTrials.gov Identifier: NCT02977819.

## INTRODUCTION

Alzheimer’s disease (AD) is characterized by the accumulation of amyloid plaques and neurofibrillary tangles, accompanied by various structural and functional brain alterations. AD development and progression is likely influenced by a combination of genetic and environmental risk factors.^1^ Among those, the apolipoprotein E allele ε4 (*APOE4*) is the major genetic risk factor for sporadic AD.^2^ Associations between *APOE4* status and brain integrity in cognitively unimpaired individuals have been inconsistent. Robust associations were found for amyloid load, showing increased amyloid deposition in *APOE4*-carriers compared to non-carriers,^3, 4^ while studies on gray matter volume (GMvol), glucose metabolism and perfusion are more discordant, showing either greater alterations,^5, 6^ greater integrity in *APOE4*-carriers,^7–9^ or no differences between groups.^3, 10, 11^

Beyond genetics, increasing evidence highlights the importance of lifestyle factors in brain integrity and cognitive performance. Cognitive activity,^12–14^ adherence to the Mediterranean diet^15–17^ and physical activity^14,18,19f^^or^ ^review^ have been associated with greater cognition and brain integrity in cognitively unimpaired older adults, although contradictory findings exist.^20^ Interestingly, *APOE4* genotype and lifestyle factors might interact on cerebral and cognitive markers of ageing and AD. More specifically, a healthy and stimulating lifestyle could particularly benefit *APOE4*-carriers.^21^ For instance, previous studies highlighted an interaction between *APOE4* status and cognitive activity on amyloid and cognition, such that higher cognitive activity was associated with lower amyloid deposition^22^ and higher cognitive scores^23^ in *APOE4*-carriers when compared to non-carriers. *APOE4*-carriers with higher adherence to the Mediterranean diet also demonstrated increased cognitive functions.^24^ Additionally, physical activity was more strongly related to increased hippocampal volume,^25^ reduced amyloid deposition^26^for review and cognitive decline^27^ in *APOE4*-carriers compared to non-carriers. However, negative findings have also been reported with no interactive effects or greater effect in *APOE*4-non-carriers for cognitive activity,^27, 28^ adherence to the Mediterranean diet^15, 29, 30^ and physical activity.^28, 31, 32^ Notably, one study found that with higher cognitive activity, *APOE4*-carriers had smaller hippocampal volumes than non-carriers.^28^ While such negative association might suggest a detrimental effect of cognitive engagement in *APOE4*-carriers, it could alternatively indicate the existence of compensation mechanisms, helping these individuals to remain cognitively normal despite brain alterations (i.e., *resilience*). The paucity of cognitive and neuroimaging outcomes concurrently prevents from interpreting the results in terms of underlying reserve mechanisms and notably to differentiate these two opposite possible interpretations.

In that context, the aim of this study was to provide a comprehensive assessment of the interplay between *APOE4* genotype and various lifestyle factors (i.e., cognitive activity, adherence to the Mediterranean diet and physical activity) on multimodal neuroimaging markers and cognition in cognitively unimpaired older adults. By combining multimodal neuroimaging and cognition, we aim to better understand the mechanisms by which lifestyle could mitigate the effect of *APOE4*. For neuroimaging, we focused on AD-sensitive markers including neocortical amyloid burden, as well as GMvol, perfusion and glucose metabolism in the Medial Temporal Lobe (MTL), known to be the first site of neurodegeneration in the course of AD^33^ and to be frequently affected in *APOE4*-carriers.^34^

Based on the existing resistance/resilience framework,^35^ we hypothesized three distinct scenarios (Figure 1). In these scenarios, *APOE4*-non-carriers are an optimal reference for healthy ageing (i.e., optimal brain and cognitive outcomes), in whom lifestyle is associated with mild to moderate increase in brain integrity and cognition. First, lifestyle could confer *resistance* to *APOE4*-carriers, allowing them to maintain brain integrity and avoid pathology. In this case, we expect healthier lifestyles in *APOE4*-carriers to be associated with greater brain integrity and cognitive function (i.e., *Resistance;* Figure 1A). Second, lifestyle could allow *APOE4*-carriers to better cope with brain aging and pathology. In this scenario, healthier lifestyles would be associated with equal or improved cognition in *APOE4*-carriers despite reduced brain integrity and/or greater pathology (i.e. *Resilience;* Figure 1B). A third hypothesis would be that lifestyle can be equally beneficial in carriers and non-carriers or cannot counteract the deleterious effect of *APOE4* carriage. In this case, no interactions between *APOE4* status and lifestyle would be evidenced in terms of both brain and cognitive outcomes (Figure 1C).

**Figure 1.**
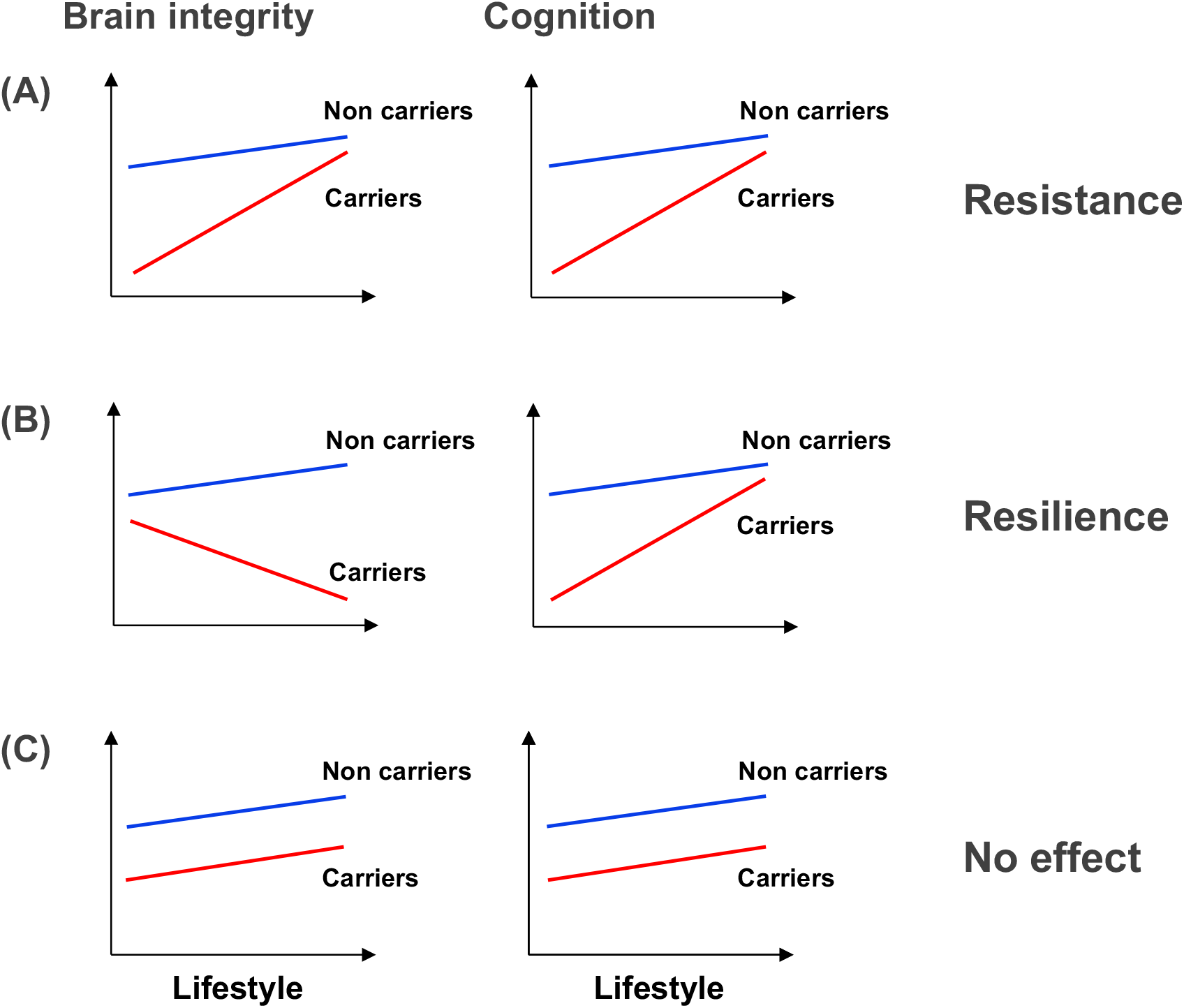
Potential mechanisms for the interaction between *APOE4* and lifestyle factors. (A) Resistance in *APOE4* carriers: lifestyle counteracts the deleterious effect of *APOE4* genotype, such that *APOE4* carriers with more active lifestyle would show higher brain integrity and cognition. (B) Resilience in *APOE4* carriers: lifestyle helps *APOE4* carriers to compensate for the presence of brain alterations and remain cognitively healthy. (C) No *APOE4* x lifestyle interaction: lifestyle does not counteract the deleterious effect of *APOE4* on brain and/or cognition. The y-axis represents brain integrity or cognition. The x-axis represents the degree of engagement in lifestyle factors. Abbreviations: APOE, apolipoprotein E gene.

## METHODS

### Participants

We included 135 cognitively unimpaired older adults from the baseline visit of the Age-Well randomized clinical trial (RCT; Medit-Ageing European Project;^36^ see Flow diagram-eFigure S1). Individuals over 65 years were recruited from the general population from November 2016 to March 2018. Participants were native French speakers, retired for at least 1 year, with at least 7 years of education, and performed within the normal range on standardized cognitive evaluation. Exclusion criteria included i) history of cerebrovascular disease, ii) presence of a chronic disease or acute unstable illness, iii) evidence of major neurological or psychiatric disorders (including drug or alcohol abuse), and iv) recent or current medication use that may interfere with cognitive functioning. Baseline data were collected between November 2016 and April 2018. Participants completed lifestyle questionnaires, and underwent *APOE4* genotyping, cognitive assessments, and multimodal neuroimaging.

### Standard Protocol Approvals, Registrations, and Patient Consents

The Age-Well RCT was approved by local ethics committee (Comité de Protection des Personnes Nord-Ouest III, Caen, France; registration number: EudraCT: 2016-002441-36; IDRCB: 2016-A01767-44; ClinicalTrials.gov Identifier: NCT02977819) and a written informed consent was obtained from each participant prior to examinations.

### Neuroimaging acquisitions

All participants were assessed on the same magnetic resonance imaging (MRI; Philips Achievia 3.0T scanner) and positron emission tomography (PET; Discovery RX VCT 64 PET-CT scanner, General Electric Healthcare) cameras in separate sessions at the Cyceron Center (Caen, France). A high-resolution T1-weighted structural image and a 3D fluid-attenuated inversion recovery image were acquired in all participants. FDG- and Florbetapir-PET scans were acquired on two separate days (n=92 and n=134, respectively). For Florbetapir-PET, each participant underwent a dual phase scan, an early acquisition beginning immediately after the injection, used as a proxy of brain perfusion (n=133), and a late acquisition (50-min post-injection) to obtain a measure of amyloid burden (n=134). Details on acquisition parameters and procedures are provided in eMethods.

### Neuroimaging processing

#### Gray matter volume, perfusion and metabolism of the Medial Temporal Lobe

The hippocampus (anterior and posterior), entorhinal, perirhinal (Brodmann areas 35 and 36) and parahippocampal cortices were automatically segmented from T1-weighted MRI with the Automatic Segmentation of Hippocampal Subfields (ASHS) software,^37^ using the ASHS-T1 atlas (ashsT1_atlas_upennpmc_07202018).^38^ For each participant, segmentation of each MTL sub-region was visually inspected. Failed segmentations were manually edited when possible, or excluded. After exclusion of failed segmentations, resulting number of participants for the GMvol of the perirhinal cortex was 131. The whole hippocampus volume was calculated as the sum of anterior and posterior hippocampus volumes while the perirhinal cortex volume was obtained by summing Brodmann areas 35 and 36 volumes. Finally, left and right volumes were averaged and normalized by the total intracranial volume (TIV, estimated using SPM12) to account for inter-individual variability in head size (normalized volume = raw volume x 1000 / TIV).

Early Florbetapir-PET was reconstructed from 1 to 6 min for a better approximation of brain perfusion. FDG and early Florbetapir-PET images were coregistered to each subject’s T1-weighted native space using rigid registration in Advanced Normalization Tools (ANTs) and then quantitatively normalized using the cerebellar cortex as a reference. ASHSs segmentations were used as regions-of-interest to extract MTL sub-regional glucose metabolism (FDG-PET) and perfusion (early Florbetapir-PET) standardized uptake value ratio (SUVr) in the resulting PET images.

#### Amyloid burden

Florbetapir-PET late acquisition images were processed as previously described, to obtain a global neocortical measure of amyloid burden.^39^ Briefly, T1-weighted MRI images were segmented using FLAIR images and spatially normalized to the Montreal Neurological Institute (MNI) space using the Statistical Parametric Mapping (SPM12) software’s multiple channels segmentation procedure (http://www.fil.ion.ucl.ac.uk/spm/software/spm12). Florbetapir-PET data were coregistered onto their corresponding T1-weighted MRI and normalized to the MNI space using the parameters from the segmentation procedure. Resulting images were quantitatively normalized using the cerebellar gray matter uptake as a reference. Global neocortical amyloid SUVr was then extracted from these images using a predetermined neocortical mask including the entire gray matter, except the cerebellum, occipital and sensory motor cortices, hippocampi, amygdala and basal nuclei.^40^

#### APOE genotyping procedure

Fasting blood samples were obtained for all participants. *APOE* genotype was identified by restriction isotyping from genomic DNA extracted from frozen leucocytes, amplified by PCR, and restricted with Hhal.^41^ *APOE4*-carriers were defined as carriers of one or two ε4 alleles. Individuals with no ε4 allele were defined as *APOE4*-non-carriers.

### Lifestyle assessment

#### Cognitive and complex mental activities

Frequency of participation in leisure cognitive activities was assessed using an adapted and French translated version of the Cognitive Activities questionnaire (CAQ)^42^ and the Lifetime of Experiences Questionnaire (LEQ).^43, 44^ In this study we focused on current lifestyle. Therefore, while both questionnaires assessed lifetime activities, only the current period of the CAQ and the late-life sub-score of the LEQ were used.

The CAQ assesses the frequency of participation in cognitive activities involved in seeking or processing information (e.g., reading books or newspapers, writing letters or emails, playing games or going to the library), relatively common and with minimal demands or requirements to participation, across different age epochs: 6, 12, 18, 40 years and the current period.^42^ For each type of activity, participants are invited to report its frequency using a 5-point response scale (1-once a year or less, 2-several times a year, 3-several times a month, 4-several times a week, 5-every day). The CAQ current period sub-score was obtained by averaging the 5 items referring to the current period, with higher scores representing a greater engagement in cognitive activities.

The LEQ assesses complex mental activity through 3 life periods: young adulthood (13–30 years), mid-life (30–65 years), and late-life (from 65 years to present date). Each life period includes a specific (e.g., education, occupational complexity) and a non-specific (e.g., visits to family or friends, music practice, writing, reading, physical activities, travelling) components, which are summed up to obtain one global score per period. Late-life LEQ (specific + non-specific) was used in the present study, with higher scores representing greater engagement in complex mental activities.

#### Adherence to the Mediterranean diet

Adherence to the Mediterranean diet was measured using the self-administered Mediterranean Diet Adherence Screener (MEDAS).^45^ This questionnaire includes 14 questions. (12 questions on food consumption frequency, and 2 questions on food intake habits) which are considered critical to assess adherence to the traditional Mediterranean diet (e.g., “do you use olive oil as the principal source of fat for cooking?”, “do you prefer to eat chicken, turkey or rabbit instead of beef, pork, hamburgers or sausages?”). One point was given to answers that were favorable to the Mediterranean diet, and 0 point to answers that were not favorable. The MEDAS score ranges from 0 to 14, with higher scores indicating higher adherence to the Mediterranean diet.

#### Physical activity

We assessed physical activity with the Modifiable Activity Questionnaire (MAQ),^46^ using the French version adapted to be self-administered.^47^ Briefly, the questionnaire evaluates the frequency and duration of leisure and work-related physical activities over the past 12 months. As all participants were retired, only the leisure activity score was considered. Participants had to select from a list all activities they did at least 10 times over the past 12 months (e.g. gardening, walking, hiking, jogging, biking), and estimate the amount of time they spent doing each activity (i.e., how many months per year, time per month and minutes each time). For each activity, the average number of hours of physical activity per week was obtained as follows: [(number of months per year) x (number of times per month) x (minutes per time) / 60 (min)] / 52 (week per year). These scores were then summed to derive a total score. The total physical activity score reflects the average hours per week of leisure-time physical activity over the past 12 months. MAQ data was missing for one participant.

#### Cognitive assessment

The neuropsychological evaluation included tests assessing global cognitive functioning, episodic memory, executive functions and attention. To obtain robust proxies of cognitive abilities and limit multiple statistical testing issues, composite scores were computed. The Preclinical Alzheimer’s Cognitive Composite 5 (PACC-5)^48^ was calculated as an index of global cognitive functioning, along with composite scores of episodic memory, executive functions and attention. Details for each score’s calculation are provided in eMethods and eTable 1.

#### Statistical analysis

First, *APOE4* carriers and non-carriers characteristics were compared using two-sample t-tests. General linear models were then used to assess i) the associations of *APOE4* and lifestyle factors separately with each neuroimaging and cognitive measure, and ii) the interactions between each lifestyle factor and *APOE4* on each neuroimaging and cognitive measure. All analyses were controlled for age, sex and education. In complementary analyses, the interactions between lifestyle factors and *APOE4* on neuroimaging or cognition were further controlled for the other lifestyle factors.

Analyses were conducted using the R software (R Core Team, 2019) and results considered significant at *p*<.05. Considering the exploratory nature of this work, no correction for multiple comparisons was applied.

The MAQ (physical activity) score was not normally distributed. Therefore, MAQ values were log-transformed after the addition of a constant of 1 to each score (as values of 0 cannot be directly log-transformed).

## RESULTS

### Participants’ characteristics

Participants’ characteristics are detailed in Table 1. The 36 *APOE4*-carriers did not differ from the 99 non-carriers in terms of age, education, female/male ratio or lifestyle.

**Table 1.** Demographics.

|  | *APOE4*<br>carriers | *APOE4* non-carriers | *p* Value |
| --- | --- | --- | --- |
| N (%) | 36 | 99 |  |
| Age, years (mean ±SD) | 69.12 (±4.13) | 69.51(±3.69) | 0.595 |
| Female/Male ratio | 18/18 | 65/34 | 0.100 |
| Education, years (mean ±SD) | 13.11 (±3.41) | 13.17 (±2.98) | 0.920 |
| Physical activity, hours per week (MAQ, mean ±SD) | 6.50 (±5.22) | 6.24 (±4.81) | 0.783 |
| Cognitive activity (CAQ, mean ±SD) | 17.56 (±3.59) | 17.53 (±3.12) | 0.962 |
| Complex mental activity (LEQ, mean ±SD) | 27.94 (±4.23) | 28.21 (±4.61) | 0.764 |
| Adherence to the Mediterranean Diet (MEDAS, mean ±SD) | 6.89 (±2.58) | 7.18(±1.99) | 0.487 |
| PACC-5 score (mean ±SD) | -0.06 (±1.14) | 0.02 (±0.95) | 0.653 |
| Episodic memory score (mean ±SD) | 0.02 (±1.11) | -0.01 (±0.97) | 0.873 |
| Executive functions score (mean ±SD) | 0.09 (±1.01) | -0.04 (±1) | 0.493 |
| Attention score (mean ±SD) | 0.08 (±1.09) | -0.03 (±0.98) | 0.566 |

P-values were obtained from two-sample t-tests. Abbreviations: SD, standard deviation; APOE, apolipoprotein E gene; PACC-5, Preclinical Alzheimer's Cognitive Composite 5.

Due to missing data (see Methods), sample size slightly varies across analysis. Final sample size per neuroimaging modality is summarized in eTable 2.

### Association of *APOE4* status with neuroimaging and cognition

*APOE4-*carriers had higher global amyloid burden compared to non-carriers (β=0.24, p=0.006). No associations were found between *APOE4* status and MTL subregions volume, metabolism, perfusion, nor with cognitive functions (all ps>0.05; eTable 3).

### Association of lifestyle factors with neuroimaging and cognition

Higher CAQ was associated with reduced GMvol in the parahippocampal cortex (β=-0.2, p=0.02). No associations were found between the CAQ and the other MTL subregions or neuroimaging modalities, nor between the LEQ, MAQ or MEDAS and neuroimaging measures (all ps>0.05; eTable 4).

Higher CAQ was associated with higher PACC-5 (β=0.23, p=0.004), executive function (β=0.24, p=0.003) and attention (β=0.25, p=0.002). Higher LEQ was associated with higher PACC-5 (β=0.2, p=0.01) and episodic memory (β=0.21, p=0.01). The MEDAS and MAQ were not associated with cognition (all ps>0.05; eTable 5).

### Interactions between *APOE4* status and lifestyle factors

Full statistics are reported in Table 2.

**Table 2.** Interactions of *APOE4* with lifestyle factors on neuroimaging values or cognitive scores.

| Lifestyle factors | GM volume |  |  |  |  |  |  |  |  |  |  |  |  |  |  |  |
| --- | --- | --- | --- | --- | --- | --- | --- | --- | --- | --- | --- | --- | --- | --- | --- | --- |
|  | Hippocampus |  |  |  | Entorhinal cortex |  |  |  | Perirhinal cortex |  |  |  | Parahippocampal cortex |  |  |  |
| | *b* | $\beta$ | *t Value* | *p Value* | *b* | $\beta$ | *t Value* | *p Value* | *b* | $\beta$ | *t Value* | *p Value* | *b* | $\beta$ | *t Value* | *p Value* |
| CAQ | -0.021 | -0.675 | -1.511 | 0.133 | 0.001 | 0.129 | 0.282 | 0.778 | -0.019 | -0.743 | -1.605 | 0.111 | -0.016 | -1.007 | -2.33 | 0.021* |
| LEQ | -0.001 | -0.035 | -0.061 | 0.951 | 0.003 | 0.607 | 1.068 | 0.288 | -0.01 | -0.595 | -1.027 | 0.306 | -0.007 | -0.721 | -1.291 | 0.199 |
| MAQ | -0.014 | -0.051 | -0.208 | 0.836 | 0.008 | 0.134 | 0.537 | 0.592 | -0.099 | -0.412 | -1.644 | 0.103 | 0.01 | 0.068 | 0.274 | 0.784 |
| MEDAS | 0.003 | 0.039 | 0.142 | 0.887 | 0.003 | 0.203 | 0.734 | 0.464 | -0.014 | -0.221 | -0.775 | 0.44 | -0.003 | -0.083 | -0.306 | 0.76 |
|  | Glucose metabolism |  |  |  |  |  |  |  |  |  |  |  |  |  |  |  |
|  | Hippocampus |  |  |  | Entorhinal cortex |  |  |  | Perirhinal cortex |  |  |  | Parahippocampal cortex |  |  |  |
| CAQ | 0.000 | 0.025 | 0.05 | 0.96 | -0.001 | -0.139 | -0.274 | 0.785 | -0.000 | -0.036 | -0.072 | 0.943 | 0.002 | 0.281 | 0.557 | 0.579 |
| LEQ | -0.002 | -0.45 | -0.671 | 0.504 | -0.002 | -0.603 | -0.889 | 0.377 | -0.004 | -0.777 | -1.158 | 0.25 | -0.006 | -1.315 | -1.992 | 0.05 |
| MAQ | 0.011 | 0.188 | 0.612 | 0.542 | 0.007 | 0.136 | 0.439 | 0.662 | 0.017 | 0.25 | 0.814 | 0.418 | -0.007 | -0.111 | -0.362 | 0.718 |
| MEDAS | 0.011 | 0.762 | 2.314 | 0.023* | 0.012 | 0.929 | 2.818 | 0.006** | 0.007 | 0.403 | 1.18 | 0.241 | 0.003 | 0.16 | 0.47 | 0.639 |
|  | Perfusion |  |  |  |  |  |  |  |  |  |  |  |  |  |  |  |
|  | Hippocampus |  |  |  | Entorhinal cortex |  |  |  | Perirhinal cortex |  |  |  | Parahippocampal cortex |  |  |  |
| CAQ | -0.003 | -0.648 | -1.406 | 0.162 | -0.006 | -1.081 | -2.357 | 0.02* | -0.006 | -1.083 | -2.351 | 0.02* | -0.003 | -0.646 | -1.455 | 0.148 |
| LEQ | -0.002 | -0.607 | -1.056 | 0.293 | -0.003 | -0.966 | -1.685 | 0.095 | -0.003 | -0.745 | -1.28 | 0.203 | -0.004 | -1.257 | -2.294 | 0.024* |
| MAQ | -0.024 | -0.512 | -2.049 | 0.043* | -0.003 | -0.069 | -0.27 | 0.788 | -0.008 | -0.151 | -0.589 | 0.557 | -0.021 | -0.418 | -1.718 | 0.088 |
| MEDAS | 0.001 | 0.059 | 0.207 | 0.836 | 0.001 | 0.102 | 0.359 | 0.72 | 0.001 | 0.104 | 0.364 | 0.716 | -0.000 | -0.016 | -0.058 | 0.954 |
|  | Amyloid load |  |  |  |  |  |  |  |  |  |  |  |  |  |  |  |
|  | Global SUVR |  |  |  |  |  |  |  |  |  |  |  |  |  |  |  |
| CAQ | 0.008 | 0.427 | 0.942 | 0.348 |  |  |  |  |  |  |  |  |  |  |  |  |
| LEQ | 0.000 | 0.001 | 0.002 | 0.998 |  |  |  |  |  |  |  |  |  |  |  |  |
| MAQ | 0.094 | 0.534 | 2.177 | 0.031* |  |  |  |  |  |  |  |  |  |  |  |  |
| MEDAS | -0.016 | -0.35 | -1.274 | 0.205 |  |  |  |  |  |  |  |  |  |  |  |  |
|  | Cognitive composite scores |  |  |  |  |  |  |  |  |  |  |  |  |  |  |  |
|  | Global cognition |  |  |  | Episodic memory |  |  |  | Executive function |  |  |  | Attention |  |  |  |
| CAQ | -0.031 | -0.25 | -0.626 | 0.533 | -0.036 | -0.283 | -0.674 | 0.502 | -0.06 | -0.483 | -1.2 | 0.232 | -0.007 | -0.055 | -0.133 | 0.895 |
| LEQ | -0.019 | -0.243 | -0.48 | 0.632 | -0.049 | -0.616 | -1.191 | 0.236 | 0.01 | 0.132 | 0.254 | 0.8 | 0.041 | 0.515 | 0.976 | 0.331 |
| MAQ | -0.057 | -0.05 | -0.22 | 0.826 | -0.152 | -0.132 | -0.568 | 0.571 | -0.181 | -0.159 | -0.692 | 0.49 | -0.087 | -0.075 | -0.326 | 0.745 |
| MEDAS | 0.082 | 0.275 | 1.099 | 0.274 | 0.008 | 0.027 | 0.106 | 0.916 | 0.09 | 0.302 | 1.196 | 0.234 | 0.156 | 0.518 | 2.026 | 0.045* |

\*P < 0.05, \*\*P < 0.01, \*\*\*P < 0.001. Abbreviations: GM, gray matter; CAQ, Cognitive Activities Questionnaire; LEQ, Lifetime of Experiences Questionnaire; MAQ, Modifiable Activity Questionnaire; MEDAS, Mediterranean Diet Adherence Screener.

### Interactions between APOE4 status and cognitive activity

*CAQ:* An interaction between *APOE4* status and the CAQ was found for GMvol in the parahippocampal cortex (β=-1.01 p=0.02), and for perfusion in the entorhinal and perirhinal cortices (β=-1.08, p=0.02 and β=-1.08, p=0.02, respectively) such that, in *APOE4*-carriers higher CAQ was associated with lower GMvol and perfusion compared to non-carriers (Figure 2A). We found no interactions between *APOE4* status and the CAQ on cognition, suggesting that the better cognition associated with higher CAQ (see paragraph 3.3) did not differ between *APOE4*-carriers and non-carriers (Figure 2B).

**Figure 2.**
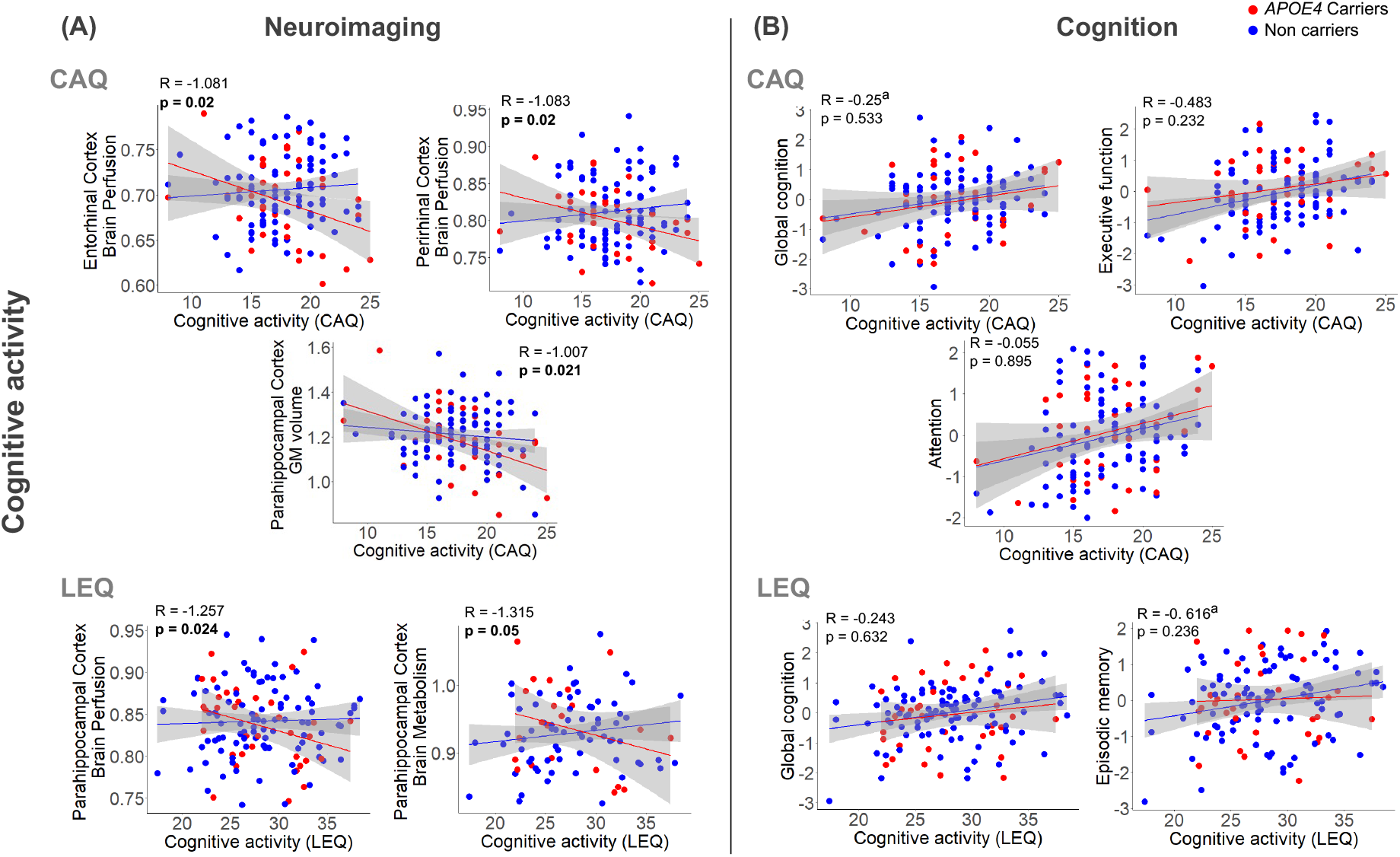
Interactive effects of cognitive activity (CAQ, LEQ) and *APOE4* on neuroimaging (A) and cognitive (B) outcomes. Statistics were obtained from general linear models controlling for age, sex and education. Raw values are plotted. Dotted lines represent confidence intervals (95%). Abbreviations: APOE, apolipoprotein E gene; CAQ, Cognitive Activities Questionnaire; LEQ, Lifetime of Experiences Questionnaire

*LEQ:* We found a significant interaction between *APOE4* status and the LEQ for perfusion in the parahippocampal cortex (β=-1.26, p=0.02), such that with higher LEQ, *APOE4*-carriers had lower perfusion compared to non-carriers (Figure 2A). A trend was found in the same direction for the parahippocampal cortex metabolism (β=-1.32, p=0.05). No interactions were found between *APOE4* status and the LEQ on cognition, suggesting that the positive association previously evidenced between the LEQ and cognition (see above) did not differ between *APOE4*-carriers and non-carriers (Figure 2B).

### Interactions between APOE4 status and adherence to the Mediterranean diet

We found an interaction between *APOE4* status and the MEDAS on hippocampal and entorhinal cortex metabolism (β=0.76, p=0.02 and β=0.93, p=0.006, respectively), such that, compared to non-carriers, *APOE4-*carriers showed a higher metabolism with higher MEDAS (Figure 3A). Similarly, an interaction between *APOE4* and the MEDAS was found on attention (β=0.52, p=0.05; Figure 3B), revealing better attention as a function of MEDAS in *APOE4*-carriers, compared to non-carriers. No interactions were found for the other neuroimaging or cognitive measures (Table 2 for details).

**Figure 3.**
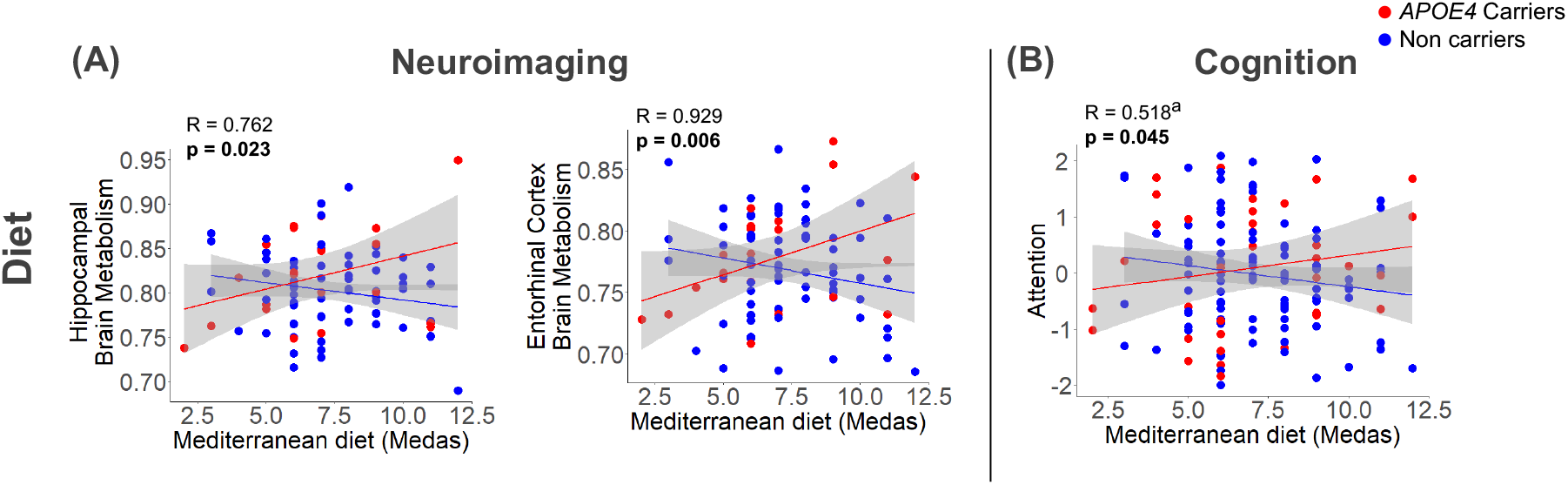
Interactive effects of Mediterranean diet adherence and *APOE4* on neuroimaging (A) and cognitive (B) outcomes. Statistics were obtained from general linear models controlling for age, sex and education. Raw values are plotted. Dotted lines represent confidence intervals (95%). Abbreviations: APOE, apolipoprotein E gene.

### Interactions between APOE4 status and physical activity

An interaction between *APOE4* status and the MAQ was found on hippocampal perfusion (β=-0.51, p=0.04; Figure 4A) and amyloid deposition (β=0.53, p=0.03; Figure 4B). The interactions were such that, compared to *APOE4-*carriers, non*-*carriers with higher MAQ scores showed higher perfusion and lower amyloid deposition. No interactions were found on the other neuroimaging or cognitive measures (Table 2).

**Figure 4.**
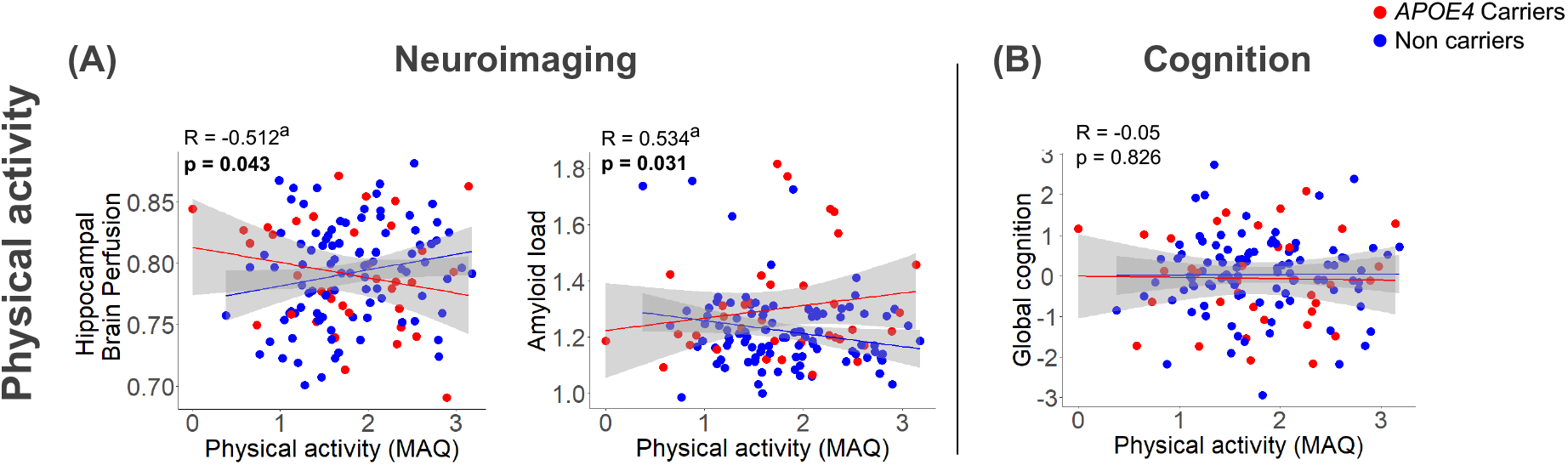
Interactive effects of physical activity and *APOE4* on neuroimaging (A) and cognitive (B) outcomes. Statistics were obtained from general linear models controlling for age, sex and education. Raw values are plotted. Dotted lines represent confidence intervals (95%). Abbreviations: APOE, apolipoprotein E gene.

### Complementary analyses

Interaction analyses were replicated while controlling for the other lifestyle factors and results remained similar, except for the interaction between *APOE4* status and the LEQ on parahippocampal cortex metabolism that became significant (β=-1.56, p=0.026), while the interaction between *APOE4* status and the MEDAS on attention was attenuated (β=0.45, p=0.07) (eTable 6).

## DICUSSION

Our study provides insights on the complex interactive effects of genetics and lifestyle factors on complementary cerebral and cognitive outcomes in cognitively unimpaired older adults. Cognitive activity was associated with lower gray matter volume and perfusion within the MTL in *APOE4* carriers only, but higher cognitive performance in both *APOE4* carriers and non-carriers. Adherence to the Mediterranean diet showed a greater positive association with both cerebral glucose metabolism and cognition (i.e. attention scores) in *APOE4* carriers when compared to non-carriers. By contrast, physical activity was associated with higher MTL perfusion and lower global amyloid load in *APOE4* non-carriers only, with no effects on cognition.

In this study, *APOE4* carriage was associated with greater amyloid deposition. This finding is consistent with a large body of evidence showing increased amyloid burden in *APOE4*-carriers.^3, 4^ However, *APOE4* was not associated with GMvol, metabolism or perfusion of the MTL, or with cognition. While deleterious effects of the *APOE4* allele on these measures have been evidenced,^5, 6, 49, 50^ the literature provides mixed results.^3, 9–11, 51, 52^ These inconsistencies could reflect the fact that, except for amyloid deposition, effects of *APOE4* in cognitively unimpaired individuals are more subtle.^3, 53^ Differences in methodology and sample characteristics across studies, including differences in lifestyle and cognitive reserve, might also explain these discrepancies.

Concerning lifestyle factors, only cognitive activity was directly associated with differences in neuroimaging markers and cognition. More particularly, higher cognitive activity was associated with lower GMvol in the parahippocampal cortex, which contrasts with previous research showing positive associations between cognitive activity and GMvol, including in the MTL.^14^ On the other hand, cognitive activity was associated with greater cognitive performance (i.e., global cognition, episodic memory, executive function and attention), in line with existing literature.^23, 54^ The association between higher cognitive activity and greater atrophy, lower perfusion and higher cognitive performance could suggest the existence of compensation mechanisms. Specifically, compensation mechanisms may be promoted by a greater engagement in cognitive activities, reflecting a *resilience* mechanism (Figure 1B).^35^ The interaction between *APOE4* status and cognitive activity suggests that this effect was mainly driven by *APOE4-*carriers.

Previous literature has highlighted interactions between *APOE4* status and cognitive activity such that cognitive activity could limit the deleterious effect of *APOE4* on AD biomarkers.^22, 55^ Conversely, a study showed that with higher cognitive activity, *APOE4*-carriers had smaller hippocampal volume than non-carriers.^28^ According to the authors, this result could reflect the existence of compensatory mechanisms in *APOE4*-carriers allowing them to maintain levels of cognitive performance comparable to non-carriers. Our findings support this view showing an interaction between *APOE4* and cognitive activity on volume and perfusion of the MTL, such that with higher engagement in cognitive activity *APOE4*-carriers had lower GMvol and perfusion, but increased cognition, similar to non-carriers. This result suggests that higher cognitive activity could promote resilience and help *APOE4*-carriers to cope with brain alterations, possibly via greater efficiency of cognitive processes^56^ (Figure 1.B).

The same pattern of interactions was replicated using two different and complementary measures of cognitive engagement (i.e., the CAQ and the LEQ), even though they concerned partially different neurodegeneration markers and cognitive domains. Specifically, *APOE4* interacted with the CAQ on parahippocampal cortex volume, entorhinal and perirhinal cortices perfusion and with the LEQ on parahippocampal cortex perfusion (and metabolism, to a lesser extent). This suggests the existence of common mechanisms, supported by different types of cognitive activities that could influence complementary markers of cognitive and brain integrity.

In the current study, adherence to the Mediterranean diet was not directly associated with any of the neuroimaging or cognitive parameters considered, in contrast with most prior research.^15–17^ However, there was an interaction with *APOE4* status on hippocampal and entorhinal cortices metabolism and attention scores. More specifically, *APOE4*-carriers with higher adherence to the Mediterranean diet showed higher metabolism and cognitive performance compared to non-carriers. These results align with previous evidence suggesting that dietary adjustments could be of particular importance to mitigate the deleterious effects of *APOE4* on brain and cognition in carriers.^57, 58^ However, prior studies provide mixed results.^15, 24, 29, 30, 59^ Methodological differences between studies (e.g., sample size, mean age, sex ratio, design) might explain these discrepancies and would require further investigations. The association of a greater adherence to Mediterranean diet with both higher MTL metabolism and attention scores in *APOE4*-carriers (in the absence of negative associations with other brain outcomes that would suggest the existence of greater brain alterations) aligns with the hypothesis of a protective effect of lifestyle that would help to limit brain changes (here neurodegeneration), in line with the concept of *resistance* (Figure 1A).^35^ Therefore, the Mediterranean diet would promote reserve in *APOE4*-carriers through mechanisms that are different from those outlined for cognitive activity, potentially by supporting brain maintenance.

Lastly, we found an interaction between *APOE4* status and physical activity on hippocampal perfusion and global amyloid burden, such that higher levels of physical activity were associated with higher perfusion in the hippocampus and lower amyloid load in non-carriers only. While this finding is in agreement with another study showing more prominent effects of exercise on AD biomarkers in *APOE4*-non-carriers,^60^ most studies report greater effects in *APOE4*-carriers.^26^ ^for^ ^review^ Previous studies reporting a beneficial effect of physical activity on hippocampal cerebral blood volume^61^ and perfusion^62^ did not investigate the modulating role of *APOE4* on these associations. Our results suggest that such effects could be driven by *APOE4*-non-carriers, in whom physical activity would support mechanisms particularly efficient to promote brain integrity.

On the other hand, physical activity was not associated with cognition, nor interacted with *APOE4* on cognition. Previous results are mixed, showing greater beneficial effect of physical activity in *APOE4*-carriers,^27^ in non-carriers,^31, 63^ in both groups,^64^ no effect,^65^ or decreased cognitive abilities in *APOE4*-carriers.^32^ These contradictory findings need to be further explored in order to clarify the mechanisms involved. Our results suggest that, contrary to cognitive activity and diet, physical activity was not associated with higher reserve in *APOE*4-carriers (Figure 1C), but was specifically associated with greater brain outcomes in non-carriers.

The strengths of this study include the assessment of multiple lifestyle factors within the same individuals, allowing investigations into the relative effect of different lifestyle factors on brain and cognitive integrity. Multimodal neuroimaging, including state-of-the-art assessment of MTL structures, combined with cognitive assessments allows a better approach to studying the reserve mechanisms underlying the moderating effect of lifestyle on *APOE4*-related changes. This study also has some limitations, including a relatively modest sample size when splitting by *APOE4* status. The use of questionnaires to assess lifestyle factors only allows the estimation of lifestyle behaviors and can be biased by individuals’ perception and memory. Nevertheless, these questionnaires are widely used and their validity and reliability have been previously reported.^42, 43, 45, 46^ Moreover, only current lifestyle was evaluated in individuals without cognitive impairments, limiting the likelihood of major recollection biases. Due to the exploratory nature of the study, no corrections were made for multiple comparisons and further replication will be needed to address the robustness of these findings. Finally, the cross-sectional design prevents evaluating the reserve mechanisms’ dynamically in *APOE4*-carriers as brain alterations progress. Longitudinal studies, including interventional studies, would enhance our understanding of the complex interactions between *APOE4* status and lifestyle.

Overall, our study demonstrates that *APOE4* genotype interacts with lifestyle to influence brain integrity and cognition with different patterns of association depending on the lifestyle factor. Our results suggest that higher cognitive activity could promote *resilience,* while higher adherence to the Mediterranean diet might instead promote *resistance* in *APOE4-*carriers. On the other hand, physical activity had a beneficial effect only in *APOE4*-non-carriers. Our findings highlight a complex interplay between genetics and lifestyle, which differentially help *APOE4*-carriers to resist or cope with brain alterations and postpone cognitive decline. In the absence of a treatment for AD, these results may deepen our understanding of how lifestyle interacts with AD genetic risk to alter the pathological phenotype and help provide lifestyle recommendations for personalized interventions to prevent or delay AD.

## Data Availability

Data is available upon request following a formal data sharing agreement and approval by the consortium and executive committee. The data sharing request form can be downloaded online: silversantestudy.eu/2020/09/25/data-sharing/

## Acknowledgments

The authors thank all the Medit-Ageing Research Group members and the Cyceron MRI-PET staff members for their help with recruitment, data acquisition, or administrative support; the EUCLID team, the sponsor (INSERM) and the participants of the study for their contribution. The full Medit-Ageing Research Group is listed in the Appendix.

---

**Appendix:**
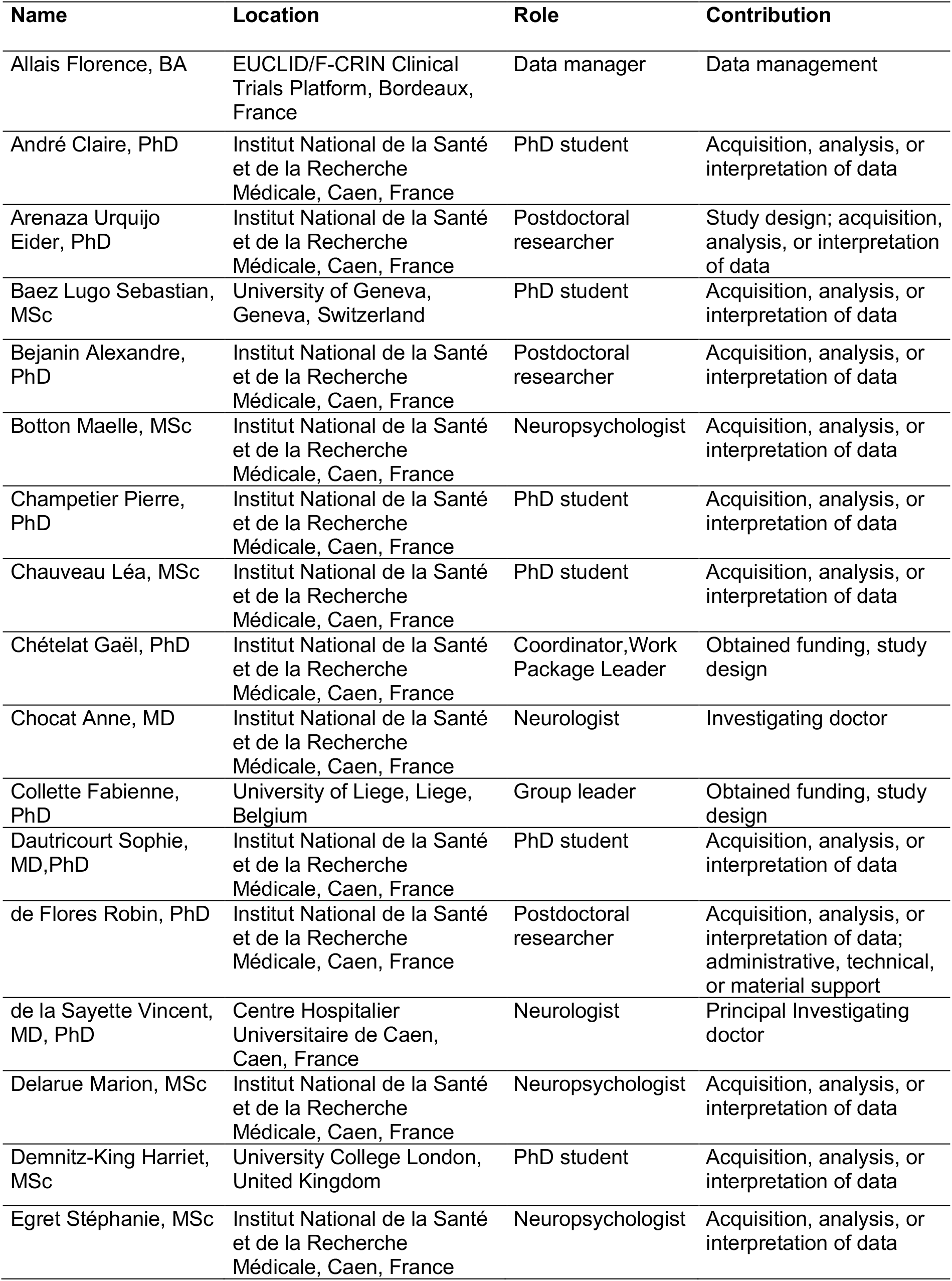
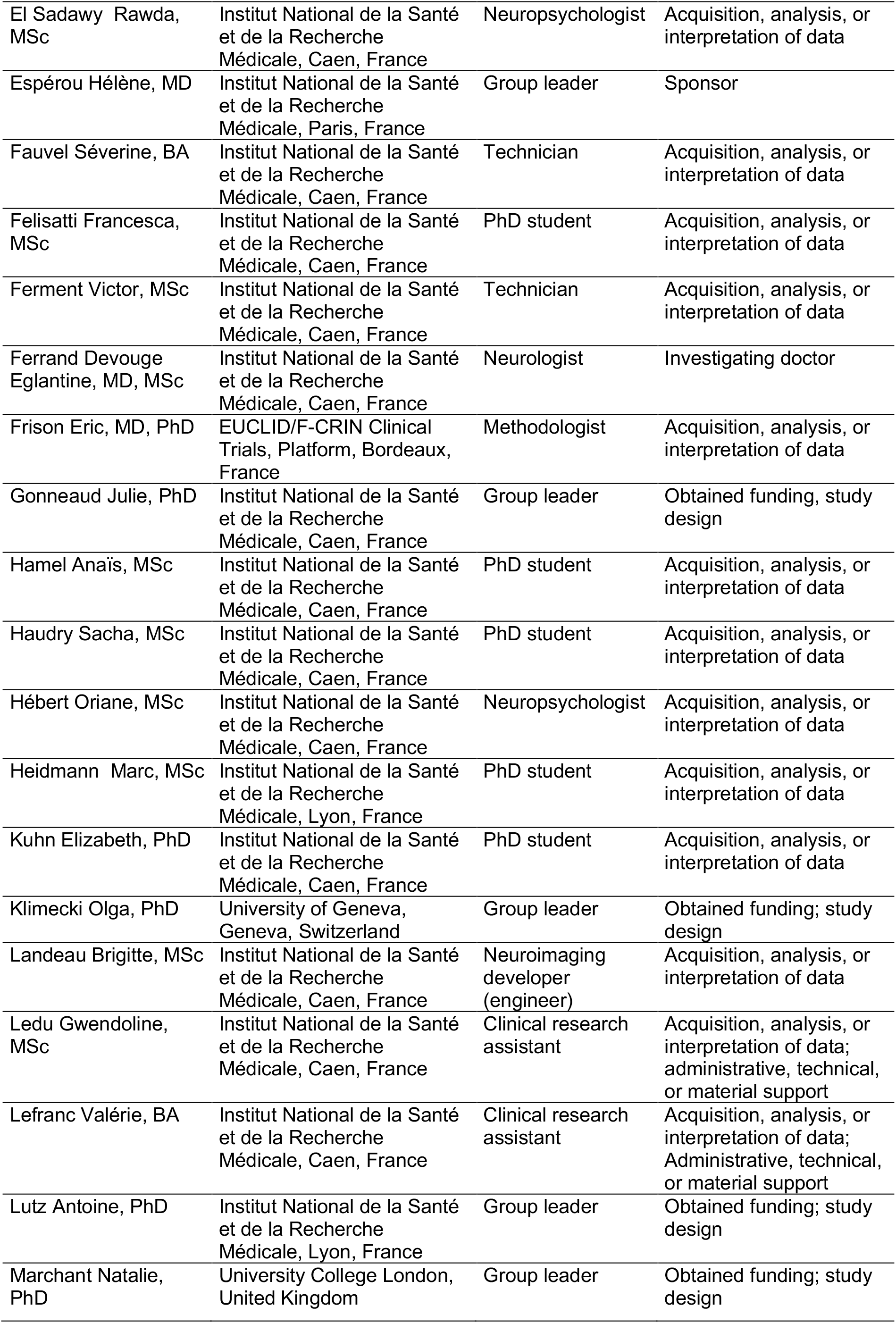
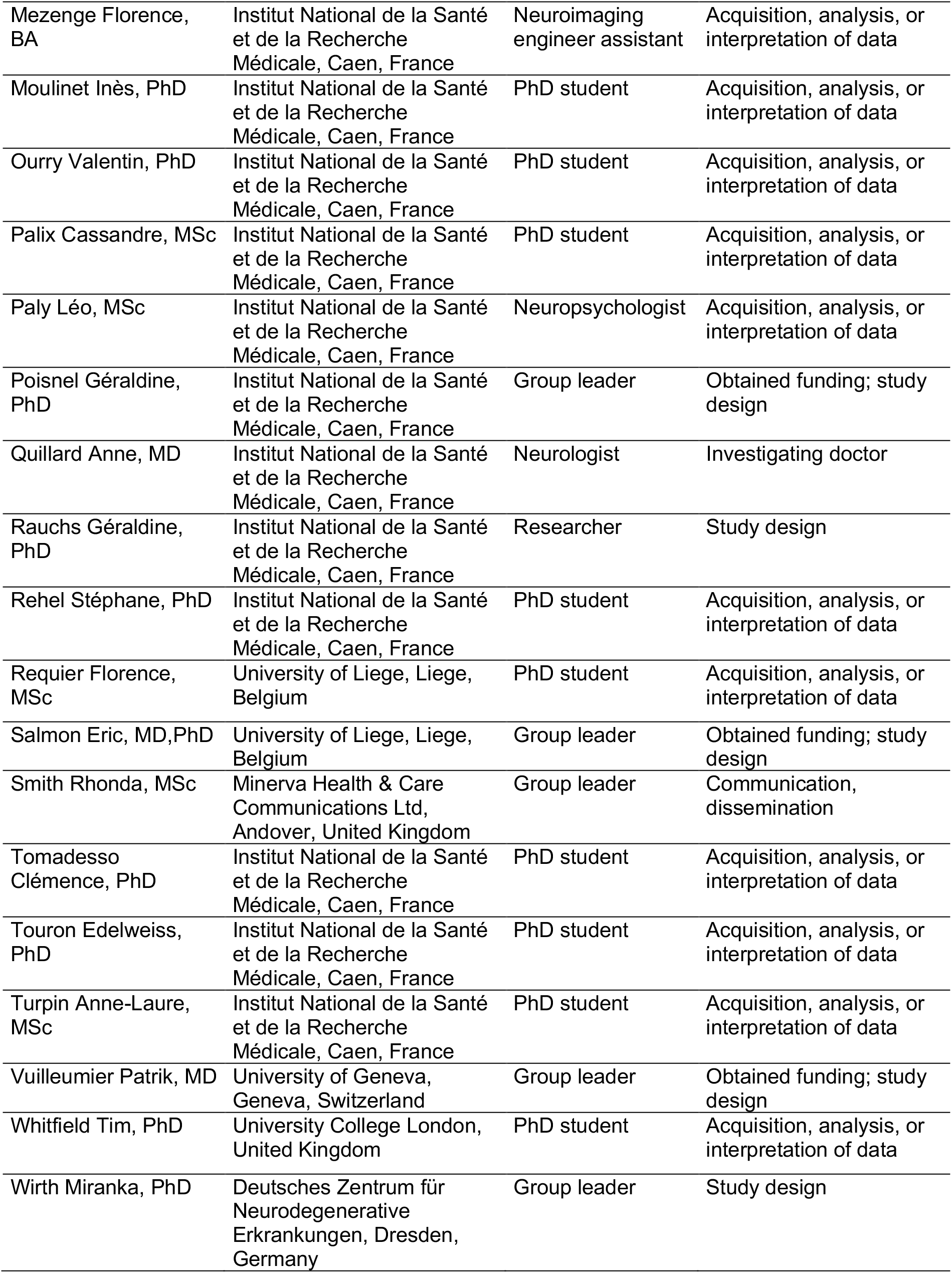
Collaborators - Medit-Ageing Research Group.

Collaborators - Medit-Ageing Research Group
| Name | Location | Role | Contribution |
| --- | --- | --- | --- |
| Allais Florence, BA | EUCLID/F-CRIN Clinical Trials Platform, Bordeaux, France | Data manager | Data management |
| André Claire, PhD | Institut National de la Santé et de la Recherche Médicale, Caen, France | PhD student | Acquisition, analysis, or interpretation of data |
| Arenaza Urquijo Eider, PhD | Institut National de la Santé et de la Recherche Médicale, Caen, France | Postdoctoral researcher | Study design; acquisition, analysis, or interpretation of data |
| Baez Lugo Sebastian, MSc | University of Geneva, Geneva, Switzerland | PhD student | Acquisition, analysis, or interpretation of data |
| Bejanin Alexandre, PhD | Institut National de la Santé et de la Recherche Médicale, Caen, France | Postdoctoral researcher | Acquisition, analysis, or interpretation of data |
| Botton Maelle, MSc | Institut National de la Santé et de la Recherche Médicale, Caen, France | Neuropsychologist | Acquisition, analysis, or interpretation of data |
| Champetier Pierre, PhD | Institut National de la Santé et de la Recherche Médicale, Caen, France | PhD student | Acquisition, analysis, or interpretation of data |
| Chauveau Léa, MSc | Institut National de la Santé et de la Recherche Médicale, Caen, France | PhD student | Acquisition, analysis, or interpretation of data |
| Chételat Gaël, PhD | Institut National de la Santé et de la Recherche Médicale, Caen, France | Coordinator, Work Package Leader | Obtained funding, study design |
| Chocat Anne, MD | Institut National de la Santé et de la Recherche Médicale, Caen, France | Neurologist | Investigating doctor |
| Collette Fabienne, PhD | University of Liege, Liege, Belgium | Group leader | Obtained funding, study design |
| Dautricourt Sophie, MD, PhD | Institut National de la Santé et de la Recherche Médicale, Caen, France | PhD student | Acquisition, analysis, or interpretation of data |
| de Flores Robin, PhD | Institut National de la Santé et de la Recherche Médicale, Caen, France | Postdoctoral researcher | Acquisition, analysis, or interpretation of data; administrative, technical, or material support |
| de la Sayette Vincent, MD, PhD | Centre Hospitalier Universitaire de Caen, Caen, France | Neurologist | Principal Investigating doctor |
| Delarue Marion, MSc | Institut National de la Santé et de la Recherche Médicale, Caen, France | Neuropsychologist | Acquisition, analysis, or interpretation of data |
| Demnitz-King Harriet, MSc | University College London, United Kingdom | PhD student | Acquisition, analysis, or interpretation of data |
| Egret Stéphanie, MSc | Institut National de la Santé et de la Recherche Médicale, Caen, France | Neuropsychologist | Acquisition, analysis, or interpretation of data |
| El Sadawy Rawda, MSc | Institut National de la Santé et de la Recherche Médicale, Caen, France | Neuropsychologist | Acquisition, analysis, or interpretation of data |
| Espérou Hélène, MD | Institut National de la Santé et de la Recherche Médicale, Paris, France | Group leader | Sponsor |
| Fauvel Séverine, BA | Institut National de la Santé et de la Recherche Médicale, Caen, France | Technician | Acquisition, analysis, or interpretation of data |
| Felisatti Francesca, MSc | Institut National de la Santé et de la Recherche Médicale, Caen, France | PhD student | Acquisition, analysis, or interpretation of data |
| Ferment Victor, MSc | Institut National de la Santé et de la Recherche Médicale, Caen, France | Technician | Acquisition, analysis, or interpretation of data |
| Ferrand Devouge Eglantine, MD, MSc | Institut National de la Santé et de la Recherche Médicale, Caen, France | Neurologist | Investigating doctor |
| Frison Eric, MD, PhD | EUCLID/F-CRIN Clinical Trials, Platform, Bordeaux, France | Methodologist | Acquisition, analysis, or interpretation of data |
| Gonneaud Julie, PhD | Institut National de la Santé et de la Recherche Médicale, Caen, France | Group leader | Obtained funding, study design |
| Hamel Anaïs, MSc | Institut National de la Santé et de la Recherche Médicale, Caen, France | PhD student | Acquisition, analysis, or interpretation of data |
| Haudry Sacha, MSc | Institut National de la Santé et de la Recherche Médicale, Caen, France | PhD student | Acquisition, analysis, or interpretation of data |
| Hébert Oriane, MSc | Institut National de la Santé et de la Recherche Médicale, Caen, France | Neuropsychologist | Acquisition, analysis, or interpretation of data |
| Heidmann Marc, MSc | Institut National de la Santé et de la Recherche Médicale, Lyon, France | PhD student | Acquisition, analysis, or interpretation of data |
| Kuhn Elizabeth, PhD | Institut National de la Santé et de la Recherche Médicale, Caen, France | PhD student | Acquisition, analysis, or interpretation of data |
| Klimecki Olga, PhD | University of Geneva, Geneva, Switzerland | Group leader | Obtained funding; study design |
| Landeau Brigitte, MSc | Institut National de la Santé et de la Recherche Médicale, Caen, France | Neuroimaging developer (engineer) | Acquisition, analysis, or interpretation of data |
| Ledu Gwendoline, MSc | Institut National de la Santé et de la Recherche Médicale, Caen, France | Clinical research assistant | Acquisition, analysis, or interpretation of data; administrative, technical, or material support |
| Lefranc Valérie, BA | Institut National de la Santé et de la Recherche Médicale, Caen, France | Clinical research assistant | Acquisition, analysis, or interpretation of data; Administrative, technical, or material support |
| Lutz Antoine, PhD | Institut National de la Santé et de la Recherche Médicale, Lyon, France | Group leader | Obtained funding; study design |
| Marchant Natalie, PhD | University College London, United Kingdom | Group leader | Obtained funding; study design |
| Mezenge Florence, BA | Institut National de la Santé et de la Recherche Médicale, Caen, France | Neuroimaging engineer assistant | Acquisition, analysis, or interpretation of data |
| Moulinet Inès, PhD | Institut National de la Santé et de la Recherche Médicale, Caen, France | PhD student | Acquisition, analysis, or interpretation of data |
| Ourry Valentin, PhD | Institut National de la Santé et de la Recherche Médicale, Caen, France | PhD student | Acquisition, analysis, or interpretation of data |
| Palix Cassandre, MSc | Institut National de la Santé et de la Recherche Médicale, Caen, France | PhD student | Acquisition, analysis, or interpretation of data |
| Paly Léo, MSc | Institut National de la Santé et de la Recherche Médicale, Caen, France | Neuropsychologist | Acquisition, analysis, or interpretation of data |
| Poisnel Géraldine, PhD | Institut National de la Santé et de la Recherche Médicale, Caen, France | Group leader | Obtained funding; study design |
| Quillard Anne, MD | Institut National de la Santé et de la Recherche Médicale, Caen, France | Neurologist | Investigating doctor |
| Rauchs Géraldine, PhD | Institut National de la Santé et de la Recherche Médicale, Caen, France | Researcher | Study design |
| Rehel Stéphane, PhD | Institut National de la Santé et de la Recherche Médicale, Caen, France | PhD student | Acquisition, analysis, or interpretation of data |
| Requier Florence, MSc | University of Liege, Liege, Belgium | PhD student | Acquisition, analysis, or interpretation of data |
| Salmon Eric, MD, PhD | University of Liege, Liege, Belgium | Group leader | Obtained funding; study design |
| Smith Rhonda, MSc | Minerva Health & Care Communications Ltd, Andover, United Kingdom | Group leader | Communication, dissemination |
| Tomadesso Clémence, PhD | Institut National de la Santé et de la Recherche Médicale, Caen, France | PhD student | Acquisition, analysis, or interpretation of data |
| Touron Edelweiss, PhD | Institut National de la Santé et de la Recherche Médicale, Caen, France | PhD student | Acquisition, analysis, or interpretation of data |
| Turpin Anne-Laure, MSc | Institut National de la Santé et de la Recherche Médicale, Caen, France | PhD student | Acquisition, analysis, or interpretation of data |
| Vuilleumier Patrik, MD | University of Geneva, Geneva, Switzerland | Group leader | Obtained funding; study design |
| Whitfield Tim, PhD | University College London, United Kingdom | PhD student | Acquisition, analysis, or interpretation of data |
| Wirth Miranka, PhD | Deutsches Zentrum für Neurodegenerative Erkrankungen, Dresden, Germany | Group leader | Study design |

