## Supplementary Material for "Interaction between *APOE4* status and lifestyle on brain and cognitive outcomes in cognitively unimpaired older adults"

**eMethods.** Neuroimaging acquisition and cognitive composite scores

**eFigure 1.** Flow diagram

**eTable 1.** Cognitive tests used to compute Cognitive composite scores

**eTable 2.** Sample size per neuroimaging modality and lifestyle factor

**eTable 3.** Multiple linear regressions between *APOE4* and neuroimaging values or cognitive scores

**eTable 4.** Multiple linear regressions between lifestyle factors and neuroimaging values

**eTable 5.** Multiple linear regressions between lifestyle factors and cognition

**eTable 6.** Interactions of *APOE4* with lifestyle factors on neuroimaging values or cognitive scores controlling for the other lifestyle factors

**eMethods**

***Neuroimaging acquisition***

A high-resolution T1-weighted structural image using a 3D fast-field echo sequence (3D-T1-FFE sagittal; repetition time (TR): 7.1ms, echo time (TE): 3.3ms, flip angle = 6°, 180 slices, field of view (FOV): 256×256 mm2, voxel size: 1×1×1mm3) and a 3D fluid-attenuated inversion recovery image (FLAIR; 3D-IR sagittal; TR: 4800ms, TE: 272ms, inversion time: 1650ms; 180 slices, FOV: 250×250mm2, voxel size: 0.98×0.98×1mm3) were acquired in all participants.

FDG- and Florbetapir-PET scans were acquired with a resolution of 3.76×3.76×4.9mm3 (FOV: 157mm). Forty-seven planes with a voxel size of 1.95×1.95×3.27mm3 were obtained. A transmission scan was performed before each PET acquisition for attenuation correction. For FDG-PET, participants (n=92) were fasted for at least 6 hours before scanning. After a 30-min resting period in a quiet and dark environment, ~180MBq of 18F-fluorodeoxyglucose were intravenously injected as a bolus and a 10-min PET acquisition scan was acquired 50 minutes after the injection. For Florbetapir-PET, each participant underwent two 10-min acquisitions, the first one beginning immediately after the intravenous injection of ~4MBq/Kg of Florbetapir, to be used as a proxy of brain perfusion (i.e. early acquisition; n=133), while the second one began 50 minutes after injection to obtain a measure of amyloid burden (n=134).

***Cognitive composite scores***

Global cognition (PACC-5):

The Preclinical Alzheimer's Cognitive Composite 5 (PACC-5) is a global cognitive composite sensitive to detecting and tracking pre-clinical AD-related decline. To calculate the PACC-5 in Age-Well, we first standardised the scores of each of its constituents: the Dementia Rating Scale-2 (total score), WAIS-IV Coding (raw score), California Verbal Learning Test (delayed free recall score), Logical Memory (delayed recall score) and Category Fluency (number of correct animals recalled in 2 minutes). We then took the unweighted average of these five z-scores, yielding the PACC-5 and re-standardised scores.

Episodic memory

A verbal episodic memory composite was created from two established measures of episodic memory: The California Verbal Learning Test, second edition (CVLT-II) and the WMS IV Logical Memory, Story B. We first standardised the scores for each of their constituent parts. For the CVLT-II this included the sum of trials 1-5, immediate free recall, delayed free recall. For Logical Memory this included immediate and delayed free recall. The unweighted average of these five z-scores yielded the Verbal Episodic Memory Composite score. Composite scores were re-standardised before analysis.

Executive function and attention

Two cognitive composite scores were computed by averaging outcomes measures of each neuropsychological test using mean and standard deviation of the entire group (Z-scores). Tasks used were the Digit span backward and Forward, Trail Making Test, Stroop test and Letter Fluency from the GREFEX battery and Digit Symbol Substitution test. The executive score was computed by averaging the standardized scores of Digit Span backward (raw score), Trail Making Test part B (response time), Stroop interference index (response time for interfering-neutral items) and Letter Fluency (raw score). The attention composite was computed by averaging the standardized scores on the Trail Making Test part A (response time), Stroop naming (response time), Digit Span forward (raw score) and Digit Symbol Substitution Test (raw score). Trail Making Test and Stroop scores were reversed so that higher individual scores and therefore higher total compound score indicate better performance. Composite scores were re-standardised (divided by standard deviation of the baseline data).

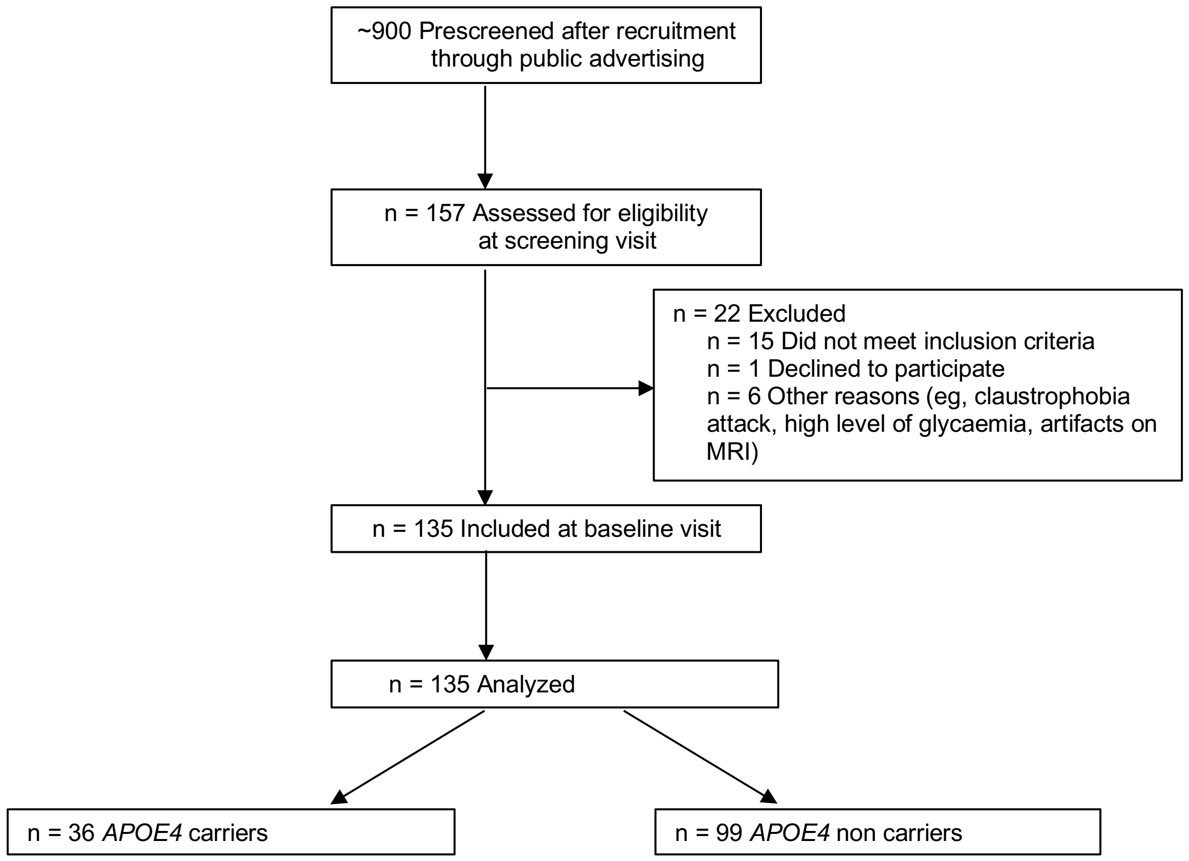


**eFigure 1. Flow diagram of the Inclusion Process.** Abbreviations: APOE, apolipoprotein E gene; FDG, $^{18}$F-fluorodeoxyglucose; PET, positron emission tomography

**eTable 1. Cognitive composite scores**

| Composite score | Tests |
|---|---|
| Global cognition (PACC-5) | Dementia Rating Scale-2 (total score)<br>WAIS-IV Coding (raw score)<br>California Verbal Learning Test (delayed free recall score)<br>Logical Memory (delayed recall score)<br>Category Fluency (number of correct animals recalled in 2 minutes) |
| Episodic memory | CVLT-II<br>WMS IV Logical Memory, Story B |
| Executive functions | Digit Span backward (raw score)<br>TMT (time TMT part-B)<br>Stroop Test (interference time)<br>Letter Fluency (raw score). |
| Attention/Processing speed | TMT (time TMT part-A)<br>Stroop Test (naming time)<br>Digit Span forward (raw score)<br>Digit Symbol Substitution Test (raw score) |

Abbreviations: PACC-5, Preclinical Alzheimer's Cognitive Composite 5, WAIS-IV, Wechsler Adult Intelligence Scale-Fourth Edition; CVLT-II, California Verbal Learning Test-Second Edition; WMS IV, Wechsler Memory Scale 4th edition; TMT, Trail Making Test.

**eTable 2. Sample size per neuroimaging modality and lifestyle factor.**

| Neuroimaging | Lifestyle factors | | | |
|---|---|---|---|---|
| | **CAQ** | **LEQ** | **Medas** | **MAQ** |
| **GM volume** | N=135 | N=135 | N=135 | N=134 |
| **Hippocampus** | N=135 | N=135 | N=135 | N=134 |
| **Entorhinal cortex** | N=135 | N=135 | N=135 | N=134 |
| **Perirhinal cortex** | N=131 | N=131 | N=131 | N=130 |
| **Parahippocampal cortex** | N=135 | N=135 | N=135 | N=134 |
| **FDG-PET** | | | | |
| **Hippocampus** | N=92 | N=92 | N=92 | N=91 |
| **Entorhinal cortex** | N=92 | N=92 | N=92 | N=91 |
| **Perirhinal cortex** | N=92 | N=92 | N=92 | N=91 |
| **Parahippocampal cortex** | N=92 | N=92 | N=92 | N=91 |
| **Early AV45-PET** | | | | |
| **Hippocampus** | N=133 | N=133 | N=133 | N=132 |
| **Entorhinal cortex** | N=133 | N=133 | N=133 | N=132 |
| **Perirhinal cortex** | N=133 | N=133 | N=133 | N=132 |
| **Parahippocampal cortex** | N=133 | N=133 | N=133 | N=132 |
| **Late AV45-PET** | | | | |
| **SUVr** | N=134 | N=134 | N=134 | N=133 |

Abbreviations: CAQ, Cognitive Activities questionnaire; LEQ, Lifetime of Experiences Questionnaire; MEDAS, Mediterranean Diet Adherence Screener; MAQ, Modifiable Activity Questionnaire; GM, gray matter; FDG, 18F-fluorodeoxyglucose; PET, positron emission tomography; SUVr, standardized uptake value ratio.

### eTable 3 Multiple linear regressions between *APOE4* and neuroimaging values or cognitive scores

| **Neuroimaging** | | | | | | | | | | | | | | | | |
|---|---|---|---|---|---|---|---|---|---|---|---|---|---|---|---|---|
| | **GM volume** | | | | | | | | | | | | | | | |
| | **Hippocampus** | | | | **Entorhinal cortex** | | | | **Perirhinal cortex** | | | | **Parahippocampal cortex** | | | |
| | ***b*** | ***ß*** | ***t* Value** | ***p* Value** | ***b*** | ***ß*** | ***t* Value** | ***p* Value** | ***b*** | ***ß*** | ***t* Value** | ***p* Value** | ***b*** | ***ß*** | ***t* Value** | ***p* Value** |
| ***APOE4* genotype** | -0.043 | -0.078 | -0.916 | 0.362 | -0.01 | -0.083 | -0.965 | 0.336 | -0.013 | -0.028 | -0.316 | 0.753 | -0.022 | -0.078 | -0.925 | 0.357 |
| | **Glucose metabolism** | | | | | | | | | | | | | | | |
| | **Hippocampus** | | | | **Entorhinal cortex** | | | | **Perirhinal cortex** | | | | **Parahippocampal cortex** | | | |
| ***APOE4* genotype** | 0.018 | 0.163 | 1.53 | 0.13 | 0.008 | 0.086 | 0.797 | 0.428 | 0.006 | 0.047 | 0.437 | 0.663 | 0.012 | 0.095 | 0.89 | 0.376 |
| | **Perfusion** | | | | | | | | | | | | | | | |
| | **Hippocampus** | | | | **Entorhinal cortex** | | | | **Perirhinal cortex** | | | | **Parahippocampal cortex** | | | |
| ***APOE4* genotype** | 0.003 | 0.029 | 0.331 | 0.741 | -0.014 | -0.148 | -1.673 | 0.097 | -0.01 | -0.1 | -1.132 | 0.26 | 0.000 | 0.004 | 0.051 | 0.96 |
| | **Amyloid load** | | | | | | | | | | | | | | | |
| | **Global SUVR** | | | | | | | | | | | | | | | |
| ***APOE4* genotype** | 0.082 | 0.237 | 2.77 | 0.006** | | | | | | | | | | | | |
| **Cognitive composite scores** | | | | | | | | | | | | | | | | |
| | **Global cognition** | | | | **Episodic memory** | | | | **Executive function** | | | | **Attention** | | | |
| ***APOE4* genotype** | -0.01 | -0.004 | -0.057 | 0.955 | 0.145 | 0.064 | 0.801 | 0.425 | 0.189 | 0.084 | 1.071 | 0.286 | 0.113 | 0.05 | 0.621 | 0.536 |

*P < 0.05, **P < 0.01, ***P < 0.001. Abbreviations: CAQ, Cognitive Activities Questionnaire; LEQ, Lifetime of Experiences Questionnaire; MAQ, Modifiable Activity Questionnaire; MEDAS, Mediterranean Diet Adherence Screener.

### eTable 4 Multiple linear regressions between lifestyle factors and neuroimaging values

| Lifestyle factors | GM volume | | | | | | | | | | | | | | | |
|---|---|---|---|---|---|---|---|---|---|---|---|---|---|---|---|---|
| | **Hippocampus** | | | | **Entorhinal cortex** | | | | **Perirhinal cortex** | | | | **Parahippocampal cortex** | | | |
| | ***b*** | ***ß*** | ***t* Value** | ***p* Value** | ***b*** | ***ß*** | ***t* Value** | ***p* Value** | ***b*** | ***ß*** | ***t* Value** | ***p* Value** | ***b*** | ***ß*** | ***t* Value** | ***p* Value** |
| **CAQ** | -0.006 | -0.072 | -0.815 | 0.417 | 0.000 | 0.011 | 0.119 | 0.906 | 0.004 | 0.063 | 0.69 | 0.492 | -0.008 | -0.198 | -2.281 | 0.024* |
| **LEQ** | -0.004 | -0.067 | -0.767 | 0.445 | 0.000 | 0.022 | 0.253 | 0.801 | 0.004 | 0.083 | 0.915 | 0.362 | -0.002 | -0.074 | -0.851 | 0.396 |
| **MAQ** | 0.056 | 0.145 | 1.733 | 0.085 | 0.007 | 0.081 | 0.948 | 0.345 | 0.046 | 0.141 | 1.627 | 0.106 | 0.013 | 0.067 | 0.798 | 0.426 |
| **MEDAS** | -0.003 | -0.029 | -0.344 | 0.732 | 0.002 | 0.074 | 0.852 | 0.396 | 0.008 | 0.087 | 0.977 | 0.33 | -0.006 | -0.098 | -1.148 | 0.253 |
| | **Glucose metabolism** | | | | | | | | | | | | | | | |
| | **Hippocampus** | | | | **Entorhinal cortex** | | | | **Perirhinal cortex** | | | | **Parahippocampal cortex** | | | |
| **CAQ** | 0.001 | 0.086 | 0.776 | 0.44 | 0.001 | 0.075 | 0.675 | 0.501 | 0.002 | 0.13 | 1.17 | 0.245 | 0.001 | 0.042 | 0.382 | 0.703 |
| **LEQ** | -0.001 | -0.048 | -0.434 | 0.665 | 0.000 | -0.002 | -0.015 | 0.988 | 0.001 | 0.115 | 1.04 | 0.301 | 0.000 | 0.000 | 0.002 | 0.999 |
| **MAQ** | 0.002 | 0.03 | 0.268 | 0.789 | 0.004 | 0.059 | 0.544 | 0.588 | 0.012 | 0.132 | 1.233 | 0.221 | 0.01 | 0.117 | 1.1 | 0.276 |
| **MEDAS** | -0.001 | -0.06 | -0.538 | 0.592 | -0.001 | -0.029 | -0.257 | 0.798 | -0.001 | -0.051 | -0.457 | 0.649 | -0.001 | -0.043 | -0.392 | 0.696 |
| | **Perfusion** | | | | | | | | | | | | | | | |
| | **Hippocampus** | | | | **Entorhinal cortex** | | | | **Perirhinal cortex** | | | | **Parahippocampal cortex** | | | |
| **CAQ** | 0.000 | 0.016 | 0.175 | 0.861 | -0.001 | -0.054 | -0.584 | 0.56 | 0.000 | 0.017 | 0.18 | 0.858 | -0.001 | -0.045 | -0.516 | 0.607 |
| **LEQ** | -0.001 | -0.112 | -1.247 | 0.215 | -0.001 | -0.154 | -1.688 | 0.094 | -0.001 | -0.095 | -1.043 | 0.299 | -0.000 | -0.042 | -0.485 | 0.628 |
| **MAQ** | 0.004 | 0.061 | 0.704 | 0.483 | 0.008 | 0.129 | 1.469 | 0.144 | 0.008 | 0.123 | 1.408 | 0.162 | 0.003 | 0.043 | 0.513 | 0.609 |
| **MEDAS** | -0.001 | -0.034 | -0.391 | 0.696 | -0.001 | -0.054 | -0.607 | 0.545 | -0.001 | -0.034 | -0.385 | 0.701 | -0.000 | -0.016 | -0.19 | 0.85 |
| | **Amyloid load** | | | | | | | | | | | | | | | |
| | **Global SUVR** | | | | | | | | | | | | | | | |
| **CAQ** | -0.003 | -0.053 | -0.582 | 0.561 | | | | | | | | | | | | |
| **LEQ** | -0.004 | -0.13 | -1.436 | 0.153 | | | | | | | | | | | | |
| **MAQ** | -0.016 | -0.068 | -0.777 | 0.439 | | | | | | | | | | | | |
| **MEDAS** | 0.004 | 0.051 | 0.573 | 0.568 | | | | | | | | | | | | |

*$P < 0.05$, **$P < 0.01$, ***$P < 0.001$. Abbreviations: GM, gray matter; CAQ, Cognitive Activities Questionnaire; LEQ, Lifetime of Experiences Questionnaire; MAQ, Modifiable Activity Questionnaire; MEDAS, Mediterranean Diet Adherence Screener.

### eTable 5 Multiple linear regressions between lifestyle factors and cognition

| Lifestyle factors | Cognitive composite scores | | | | | | | | | | | | | | | |
|---|---|---|---|---|---|---|---|---|---|---|---|---|---|---|---|---|
| | **Global cognition** | | | | **Episodic memory** | | | | **Executive function** | | | | **Attention** | | | |
| | ***b*** | ***ß*** | ***t Value*** | ***p Value*** | ***b*** | ***ß*** | ***t Value*** | ***p Value*** | ***b*** | ***ß*** | ***t Value*** | ***p Value*** | ***b*** | ***ß*** | ***t Value*** | ***p Value*** |
| **CAQ** | 0.071 | 0.231 | 2.945 | 0.004** | 0.045 | 0.145 | 1.755 | 0.082 | 0.075 | 0.243 | 3.043 | 0.003** | 0.078 | 0.251 | 3.104 | 0.002** |
| **LEQ** | 0.043 | 0.196 | 2.497 | 0.014* | 0.047 | 0.213 | 2.63 | 0.01** | 0.031 | 0.139 | 1.718 | 0.088 | 0.03 | 0.134 | 1.63 | 0.105 |
| **MAQ** | -0.003 | -0.007 | -0.084 | 0.933 | 0.002 | 0.001 | 0.014 | 0.989 | -0.14 | -0.09 | -1.155 | 0.25 | -0.183 | -0.117 | -1.488 | 0.139 |
| **MEDAS** | 0.003 | 0.01 | 0.095 | 0.924 | 0.027 | 0.058 | 0.718 | 0.474 | -0.04 | -0.087 | -1.095 | 0.275 | -0.021 | -0.046 | -0.566 | 0.572 |

*P < 0.05, **P < 0.01, ***P < 0.001. Abbreviations: CAQ, Cognitive Activities Questionnaire; LEQ, Lifetime of Experiences Questionnaire; MAQ, Modifiable Activity Questionnaire; MEDAS, Mediterranean Diet Adherence Screener.

**eTable 6 Interactions of *APOE4* with lifestyle factors on neuroimaging values or cognitive scores controlling for the other lifestyle factors**

| **Lifestyle factors** | **GM volume** | | | | | | | | | | | | | | | |
|---|---|---|---|---|---|---|---|---|---|---|---|---|---|---|---|---|
| | **Hippocampus** | | | | **Entorhinal cortex** | | | | **Perirhinal cortex** | | | | **Parahippocampal cortex** | | | |
| | ***b*** | ***ß*** | ***t Value*** | ***p Value*** | ***b*** | ***ß*** | ***t Value*** | ***p Value*** | ***b*** | ***ß*** | ***t Value*** | ***p Value*** | ***b*** | ***ß*** | ***t Value*** | ***p Value*** |
| **CAQ** | -0.022 | -0.727 | -1.601 | 0.112 | 0.000 | 0.048 | 0.102 | 0.919 | -0.023 | -0.896 | -1.904 | 0.059 | -0.016 | -1.013 | -2.288 | 0.024* |
| **LEQ** | -0.001 | -0.074 | -0.126 | 0.9 | 0.002 | 0.539 | 0.904 | 0.368 | -0.014 | -0.856 | -1.424 | 0.157 | -0.005 | -0.515 | -0.893 | 0.374 |
| **MAQ** | -0.008 | -0.028 | -0.114 | 0.909 | 0.008 | 0.124 | 0.49 | 0.625 | -0.103 | -0.432 | -1.703 | 0.091 | 0.016 | 0.113 | 0.464 | 0.644 |
| **MEDAS** | 0.004 | 0.048 | 0.172 | 0.864 | 0.003 | 0.195 | 0.69 | 0.491 | -0.015 | -0.244 | -0.841 | 0.402 | -0.001 | -0.02 | -0.074 | 0.942 |
| | **Glucose metabolism** | | | | | | | | | | | | | | | |
| | **Hippocampus** | | | | **Entorhinal cortex** | | | | **Perirhinal cortex** | | | | **Parahippocampal cortex** | | | |
| **CAQ** | 0.000 | 0.069 | 0.134 | 0.894 | -0.001 | -0.14 | -0.267 | 0.79 | -0.001 | -0.085 | -0.166 | 0.869 | 0.002 | 0.271 | 0.526 | 0.6 |
| **LEQ** | -0.003 | -0.657 | -0.934 | 0.353 | -0.003 | -0.807 | -1.131 | 0.261 | -0.005 | -1.034 | -1.489 | 0.14 | -0.007 | -1.563 | -2.273 | 0.026* |
| **MAQ** | 0.01 | 0.182 | 0.586 | 0.56 | 0.007 | 0.128 | 0.405 | 0.686 | 0.015 | 0.231 | 0.746 | 0.458 | -0.007 | -0.11 | -0.352 | 0.726 |
| **MEDAS** | 0.011 | 0.725 | 2.147 | 0.035* | 0.012 | 0.905 | 2.677 | 0.009** | 0.006 | 0.359 | 1.046 | 0.299 | 0.002 | 0.123 | 0.355 | 0.723 |
| | **Perfusion** | | | | | | | | | | | | | | | |
| | **Hippocampus** | | | | **Entorhinal cortex** | | | | **Perirhinal cortex** | | | | **Parahippocampal cortex** | | | |
| **CAQ** | -0.003 | -0.64 | -1.364 | 0.175 | -0.006 | -1.096 | -2.372 | 0.019* | -0.006 | -1.123 | -2.41 | 0.017* | -0.004 | -0.671 | -1.469 | 0.144 |
| **LEQ** | -0.002 | -0.757 | -1.262 | 0.209 | -0.004 | -1.113 | -1.87 | 0.064 | -0.003 | -0.93 | -1.543 | 0.125 | -0.005 | -1.329 | -2.307 | 0.023* |
| **MAQ** | -0.024 | -0.508 | -2.02 | 0.046* | -0.002 | -0.044 | -0.171 | 0.865 | -0.007 | -0.14 | -0.543 | 0.588 | -0.02 | -0.41 | -1.662 | 0.099 |
| **MEDAS** | 0.000 | 0.013 | 0.045 | 0.964 | 0.001 | 0.078 | 0.273 | 0.785 | 0.001 | 0.054 | 0.186 | 0.853 | -0.000 | -0.015 | -0.056 | 0.956 |
| | **Amyloid load** | | | | | | | | | | | | | | | |
| | **Global SUVR** | | | | | | | | | | | | | | | |
| **CAQ** | 0.009 | 0.454 | 0.985 | 0.327 | | | | | | | | | | | | |
| **LEQ** | -0.001 | -0.048 | -0.081 | 0.936 | | | | | | | | | | | | |
| **MAQ** | 0.093 | 0.528 | 2.15 | 0.034* | | | | | | | | | | | | |
| **MEDAS** | -0.016 | -0.349 | -1.256 | 0.212 | | | | | | | | | | | | |
| | **Cognitive composite scores** | | | | | | | | | | | | | | | |
| | **Global cognition** | | | | **Episodic memory** | | | | **Executive function** | | | | **Attention** | | | |
| **CAQ** | -0.034 | -0.27 | -0.667 | 0.506 | -0.047 | -0.377 | -0.889 | 0.376 | -0.051 | -0.407 | -0.999 | 0.32 | 0.003 | 0.023 | 0.055 | 0.956 |
| **LEQ** | -0.032 | -0.4 | -0.773 | 0.441 | -0.059 | -0.739 | -1.369 | 0.174 | 0.004 | 0.045 | 0.086 | 0.931 | 0.037 | 0.46 | 0.867 | 0.388 |
| **MAQ** | -0.089 | -0.078 | -0.357 | 0.722 | -0.189 | -0.165 | -0.719 | 0.473 | -0.202 | -0.177 | -0.801 | 0.425 | -0.118 | -0.103 | -0.458 | 0.647 |
| **MEDAS** | 0.063 | 0.211 | 0.866 | 0.388 | -0.001 | -0.005 | -0.018 | 0.986 | 0.072 | 0.242 | 0.983 | 0.328 | 0.135 | 0.448 | 1.81 | 0.073 |

*P < 0.05, **P < 0.01, ***P < 0.001. Abbreviations: GM, gray matter; CAQ, Cognitive Activities Questionnaire; LEQ, Lifetime of Experiences Questionnaire; MAQ, Modifiable Activity Questionnaire; MEDAS, Mediterranean Diet Adherence Screener.
